# Manuscript title: Loss in the expansion of SARS-CoV-2 specific immunity is a key risk factor in fatal patients with COVID-19

**DOI:** 10.1101/2020.07.29.20164681

**Authors:** Qiang Zeng, Gang Huang, Yong-zhe Li, Guoqiang Xu, Sheng-yong Dong, Tian-yu Zhong, Zong-tao Chen, Yang Xu

**Author notes:** **Contact information for the corresponding author:** Yang Xu, MD, PhD, Director, Laboratory of Special Diagnosis, Shanghai University of Medicine and Health Sciences, Shanghai 201318, China.

## Abstract

Knowledge of the dynamic immunological characteristics of patients with coronavirus disease 2019 (COVID-19) is essential for clinicians to understand the progression of the disease. Our data showed that the immune system and function gradually remodeled and declined with age, starting from age 16 until age 91 in 25,239 healthy controls. An analysis of the relationship between the number of lymphocytes and age revealed that the lymphocyte and subset counts tended to decline with age significantly. Severe acute respiratory syndrome coronavirus 2 (SARS-CoV-2)-specific immunity declined with age and was associated with survival time in fatal cases. Loss in the expansion of SARS-CoV-2-specific immunity could be expanded *in vitro*. The concurrent decline in SARS-CoV-2-specific cellular and humoral immunities and prolonged SARS-CoV-2 exposure predicted fatal outcomes. Our findings provide a basis for further analysis of SARS-CoV-2-specific immunity and understanding of the pathogenesis of fatal COVID-19 cases.

Highlights
- **The immune system and function gradually remodeled and declined with age**.
- **SARS-CoV-2-specific immunity declined with age in fatal cases**.
- **SARS-CoV-2-specific immunity was associated with survival time in fatal cases**.
- **Loss in the expansion of SARS-CoV-2-specific immunity could be expanded *in vitro***.
- **A concurrent decline in SARS-CoV-2-specific cellular and humoral immunities and prolonged SARS-CoV-2 exposure predicted fatal outcomes**.

## INTRODUCTION

Coronavirus disease 2019 (COVID-19) pandemic leads to severe illness, life-threatening complications, and death, especially in high-risk groups such as elderly people and individuals with comorbidities^1-7^. However, information on immunological characteristics in the assessment of COVID-19 for frontline clinicians is limited. The objective of this study is to explore the dynamic immunological characteristics as the disease progresses.

In January, 2020, the severe acute respiratory syndrome coronavirus 2 (SARS-CoV-2) was identified in samples of bronchoalveolar lavage fluid from patients in Wuhan, China, and was confirmed as the cause of the SARS-CoV-2 pneumonia^1,2^. Full-genome sequencing and phylogenic analysis indicated that SARS-CoV-2 is a distinct clade from the betacoronaviruses associated with human SARS and Middle East respiratory syndrome (MERS)^1,2^

COVID-19 has spread rapidly since it was first identified in Wuhan and has been shown to have a wide spectrum of severity. Recently, a report shows that SARS-CoV and SARS-CoV-2 shared the same functional host-cell receptor, angiotensin-converting enzyme 2 (ACE2)^8^. Furthermore, SARS-CoV-2 binds to ACE2 receptors in 10-20 fold higher affinity than SARS-CoV binds to the same receptors^8^. The receptor binding domain is recognized by the extracellular peptidase domain of ACE2 mainly through polar residues, which provide important insights into the molecular basis for coronavirus recognition and infection^9^. Since ACE2 receptors on lung alveolar epithelial cells and enterocytes of the small intestine are highly expressed, lung alveolar epithelial cells or enterocytes of the small intestine may be an important source of infection^10^.

According to World Health Organization (WHO) interim guidance on January 12, 2020, SARS-CoV-2 infection is classified as asymptomatic, mild, severe, and critical cases of pneumonia (acute respiratory distress syndrome [ARDS], sepsis, septic shock). Severe cases of pneumonia are defined as patients with respiratory rate > 30 breaths/min, severe respiratory distress, or peripheral capillary oxygen saturation < 90% on room air^11^. Asymptomatic cases have been reported in China and Germany^12,13^. Huang et al.^2^ first reported 41 cases of SARS-CoV-2 pneumonia in which most patients had a history of exposure to Huanan Seafood Wholesale Market. Organ dysfunction (shock, ARDS, acute cardiac injury, and acute kidney injury, etc.) and death can occur in severe or critical cases. Guan et al.^3^ reported findings from 1,099 cases of SARS-CoV-2 pneumonia and the results suggested that the SARS-CoV-2 infection clustered within groups of people in close contact, and was more likely to affect the old with comorbidities. However, there is no pre-existing antibody-mediated immunity to SARS-CoV-2 at the population level, leading to more than half million deaths and 10 millions of infections which have been reported worldwide as of June 29, 2020^14^

The disease severity is associated with immune dysregulation, for example, dysregulation of T cells, or associated with macrophage activation syndrome^15-21^. Grifoni et al.^22^ reported SARS-CoV-2-specific CD4 and CD8 T cells responses in 20 recovery cases of COVID-19. Using multiple experimental approaches, SARS-CoV-2-specific CD4 T cell and antibody responses were observed in all COVID-19 patients and SARS-CoV-2-specific CD8 T cell responses were seen in most COVID-19 patients. All of the patients had SARS-CoV-2-specific CD4 T cells that recognized the virus’s spike protein, and 70% of them had SARS-CoV-2-specific CD8 T cells that responded to the same protein. The identification of strong T cell responses in recovered COVID-19 patients promotes further study in designing vaccines to induce T cell responses. Ni et al.^18^ also observed SARS-CoV-2-specific humoral and cellular immunity in 14 convalescent patients. However, to the best of our knowledge, SARS-CoV-2-specific CD4 and CD8 T cell responses in fatal cases of COVID-19 were not reported. Furthermore, understanding the key immune mechanisms to recover from novel SARS-CoV-2 exposure is an urgent need.

It is important to understand the key risk factors of critical patients who died. We hypothesize that lethal COVID-19 disease would be associated with immune dysregulation and loss expansion of SARS-CoV-2-specific immunity. Here, we analyzed dynamic immunological characteristics in fatal COVID-19 patients in a unique longitudinal cohort of samples.

## RESULTS

### The immune system and function gradually remodeled and declined with age in 25,239 healthy controls

The numbers of lymphocytes and subset counts, except for natural killer (NK) cells, tended to decline with age significantly (Spearman *R* = - [range, 0.9048-1], *P* < 0.001, Figure 1A-E and Table S2). However, the relationship between the number of NK cells and age only showed a tendency of the NK cells to increase in number with age (Spearman R = 0.6429, *P* = 0.0962, Figure 1F and Table S2).

**Figure 1.**
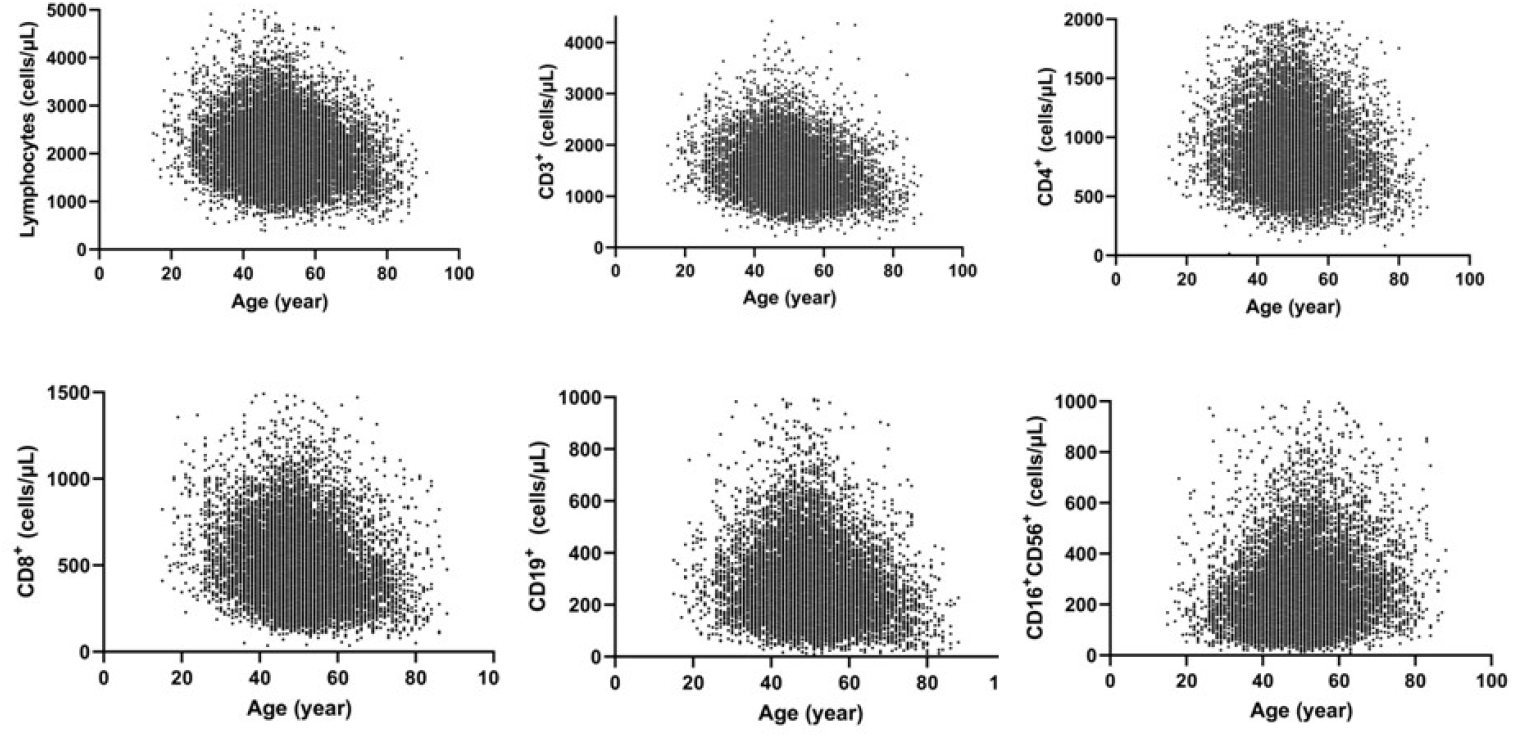
Dynamic distribution of lymphocytes and subsets according to age among the 25,239 healthy controls. We enrolled laboratory data from 25,239 health controls in November 2017 and November 2019 from health check up before the COVID-19 pandemic.

### SARS-CoV-2-speciflc immunity declined with age in fatal COVID-19 cases

The SARS-CoV-2-specific immunity tended to decline with age (Figure 2A-C). However, the virus load tended to increase with age (Figure 2D).

**Figure 2.**
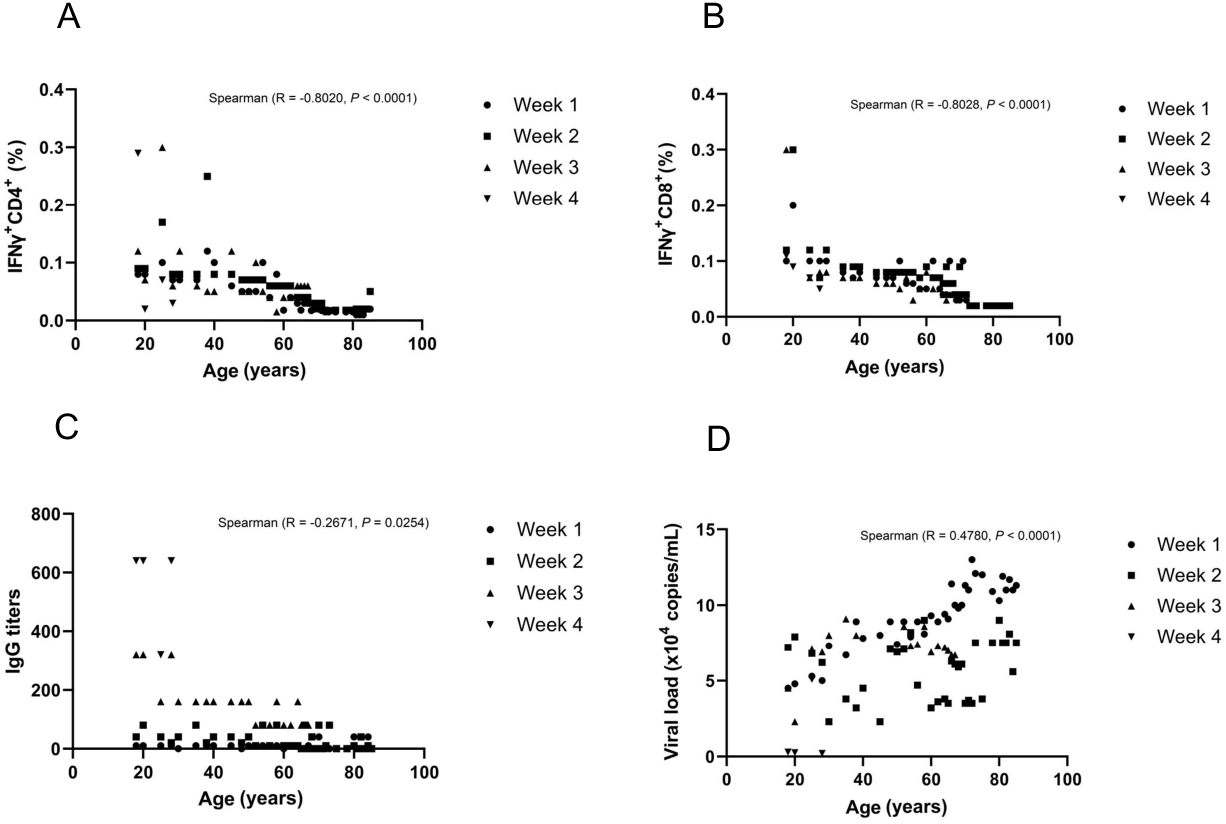
Dynamic distribution of severe acute respiratory syndrome coronavirus 2 (SARS-CoV-2)-specific immunity and viral load according to age among the fatal cases.

The ability to expand SARS-CoV-2-specific interferon gamma (IFNγ)+CD4+ T cells (Spearman *R* = −0.8020, *P*< 0.0001, Figure 2A) and IFNγ+CD8+ T cells (Spearman *R* = −0.8028, *P*< 0.0001, Figure 2B) was significantly inversely related to age. This suggests that the elderly had low expansion ability of SARS-CoV-2-specific T-cell immunity, which led to the faster progression of the disease to a severe illness, an important risk factor of mortality in the elderly. SARS-CoV-2-specific humoral immunity (Spearman *R* = −0.2671, *P* = 0.0254, Figure 2C) was also inversely related to age. However, this relationship was weaker than that of SARS-CoV-2-specific T-cell immunity with age. The SARS-CoV-2 viral load was directly and significantly proportional to age (Spearman *R* = 0.4780, *P*< 0.0001, Figure 2D). This suggests that the elderly easily contracted the infection or that the patients had a higher viral load in the body.

### SARS-CoV-2-specific immunity is associated with survival time in fatal COVID-19 cases

As the survival time increased, the SARS-CoV-2-specific immunity tended to increase. However, the relationship between virus load and survival time showed that the virus load tended to decrease as the survival time increased (Figure 3).

**Figure 3.**
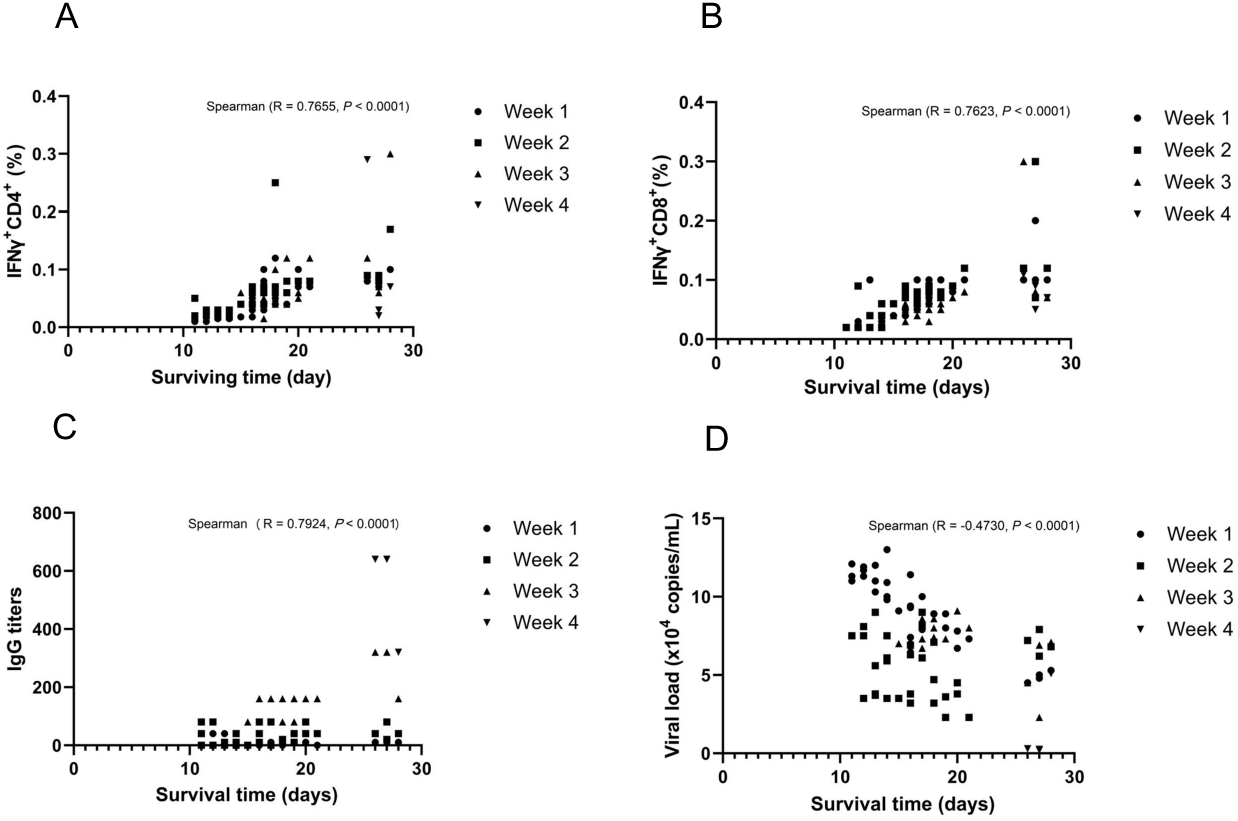
Dynamic distribution of severe acute respiratory syndrome coronavirus2 (SARS-CoV-2)-specific immunity and viral load according to survival time among the fatal cases.

The correlations between SARS-CoV-2-specific IFNγ+CD4+ T cells (Spearman *R* = 0.7655, *P*< 0.0001, Figure 3A) and IFNγ+CD8+ T cells (Spearman *R* = 0.7623, *P*< 0.0001, Figure 3B) and survival time were relatively strong and significant, suggesting that the patient’s survival time depended on the expansion ability of SARS-CoV-2-specific T-cell immunity. The aggregated data for 4 weeks showed that SARS-CoV-2-specific humoral immunity was significantly related to survival time (Spearman *R* = 0.7924, *P*< 0.0001, Figure 3C). The SARS-CoV-2 viral load was inversely proportional to survival time (Spearman *R* = −0.4730, *P*< 0.0001, Figure 3D). This suggests that the higher the viral load of SARS-CoV-2, the shorter the survival time of the patient. Therefore, controlling the replication of the virus might prolong the survival time of patients.

We found no significant difference in the frequency of SARS-CoV-2-specific IFNγ+CD4+ T cells (Figure 4A) between the first and second weeks. However, significant differences were observed between the first and third or fourth weeks, possibly indicating that SARS-CoV-2-specific IFNγ+CD4+ T cells began to expand in the third week significantly (Figure 4A). No significant difference in the frequency of SARS-CoV-2 specific IFNγ+CD8+ T cells was found between the first and second, third, or fourth weeks, indicating that the expansion of the specific CD8 T cells was lost (Figure 4B). A significant difference in the titer of the specific IgG antibodies was found between the first and second weeks. Compared with that at week 1, the IgG titer in weeks 2–4 continued to show significant differences, which suggests that the SARS-CoV-2-specific antibodies began to expand in the second week, but the antibody titer was low (Figure 4C). Significant differences were found between the viral loads from weeks 1 to 4 (Figure 4D), indicating that the SARS-CoV-2-specific immunity could potentially clear the SARS-CoV-2 replication in fatal cases. However, the SARS-CoV-2-specific immunity was unable to clear all SARS-CoV-2 replications in the fatal cases. The mean viral load was 6.7 × 10^4^ copies/mL at week 3, indicating the presence of persistent SARS-CoV-2 infection.

**Figure 4.**
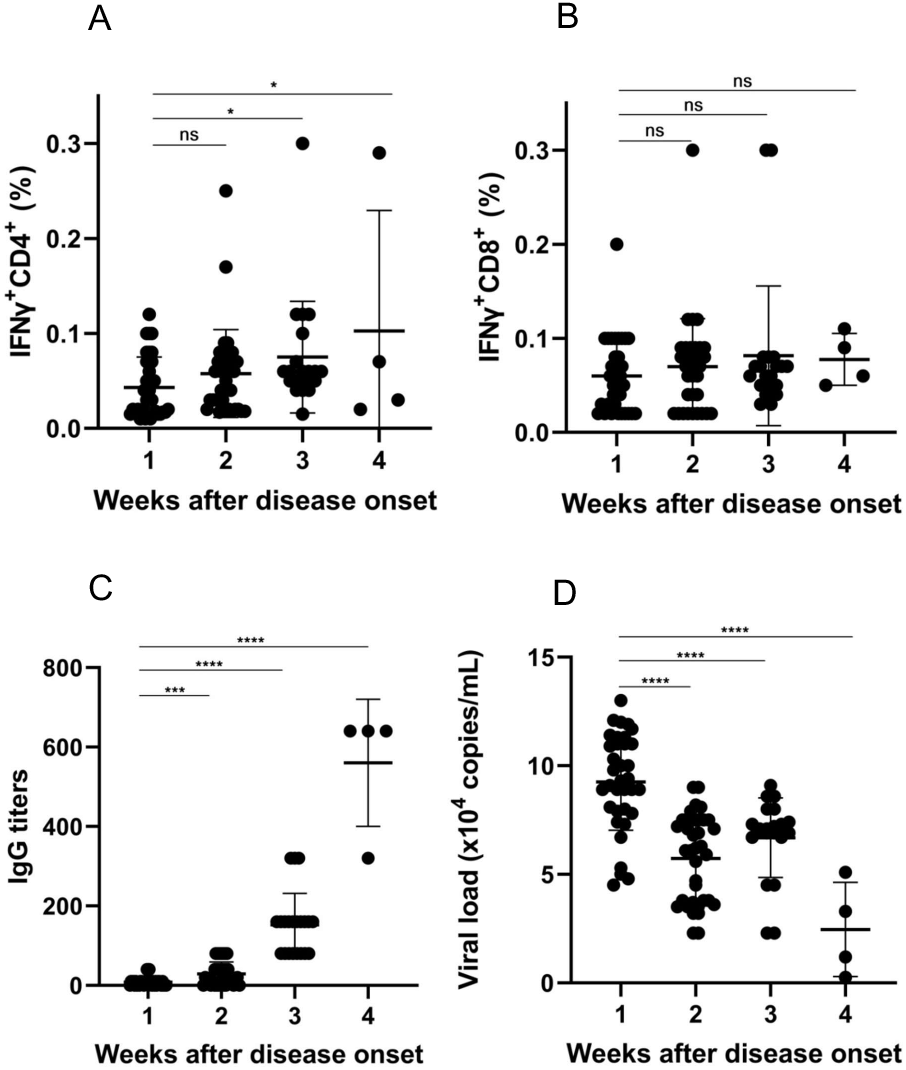
Dynamic characteristics of severe acute respiratory syndrome coronavirus 2 (SARS-CoV-2)-specific immunity and viral load in the fatal cases. ns, *P*> 0.05; * *P*< 0.05; ** *P*< 0.01; *** *P*< 0.001; **** *P*< 0.0001.

### The loss in the expansion of SARS-CoV-2-specific immunity in the fatal cases could be expanded *in vitro*

From our results, we can infer that the key factor was loss in the expansion of SARS-CoV-2-specific humoral and cellular immunities, causing the virus to remain in the body in fatal cases. As the expansion of SARS-CoV-2-specific immunity started from the first 2–3 weeks, we chose peripheral blood mononuclear cells (PMBCs) in the third week for the expansion experiment *in vitro*. Our data showed that these PBMCs could be expanded *in vitro*. After the expansion, SARS-CoV-2-specific IFNγ+CD4+ T cells and IFNγ+CD8+ T cells increased significantly (Figure 5). This suggests that the SARS-CoV-2-specific immunity lost its expansion in the 21 fatal cases at week 3, but the expansion of the SARS-CoV-2-specific cellular immunity was recovered in the *in vitro* experiment (Figure 5).

**Figure 5.**
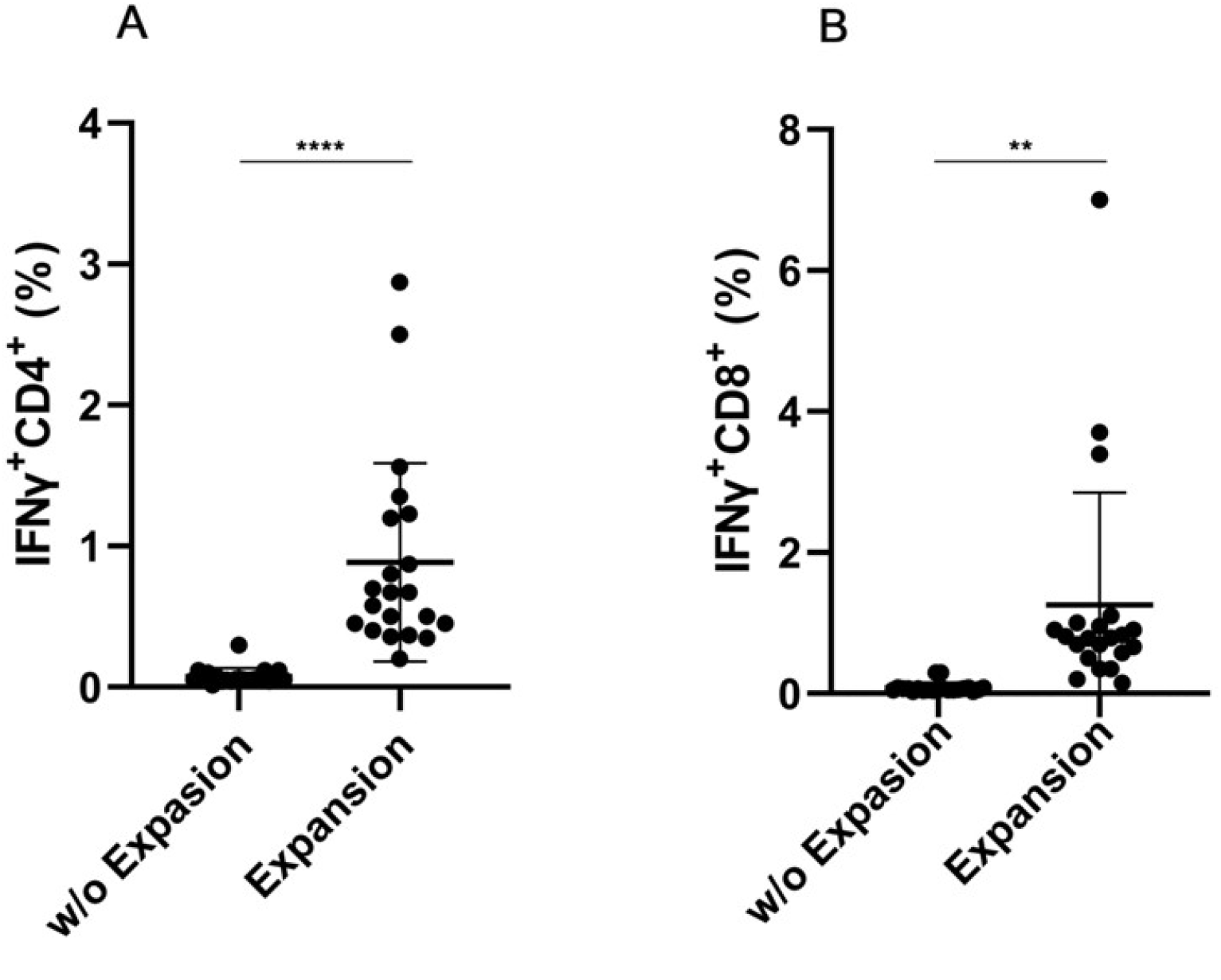
Peripheral blood mononuclear cells (PBMCs) of the 21 fatal cases at week 3 stimulated with severe acute respiratory syndromecoronavirus2 (SARS-CoV-2) spike glycoprotein peptide pools (SPs). The graph showed the percentage of SARS-CoV-2 specific IFNγ+CD4+T cells (A) and SARS-CoV-2 specific IFNγ+CD8+ T cells (B) reactive to the SPs directly *ex vivo* (w/o expansion) and after *in vitro* expansion (expansion). **, *P*=0.0017; ****, *P*<0.0001.

### A concurrent decline in SARS-CoV-2-specific cellular and humoral immunities and prolonged SARS-CoV-2 exposure predicted fatal outcomes

Significant correlations were observed between SARS-CoV-2-specific IFNγ+CD4+ T cells (Spearman *R* = 0.4537, *P*< 0.0001, Figure 6A) and IFNγ+CD8+ T cells (Spearman *R* = 0.3721, *P* = 0.0002, Figure 6B) and SARS-CoV-2-specific humoral immunity.

SARS-CoV-2-specific IFNγ+CD4+ T cells (Spearman *R* = −0.4784, *P*< 0.0001, Figure 7A) and IFNγ+CD8+ T cells (Spearman *R* = −0.4609, *P*< 0.0001, Figure 7B) inversely correlated significantly with the SARS-CoV-2 viral load, which suggests that the SARS-CoV-2-specific T-cell immunity plays a leading role in viral clearance.

**Figure 6.**
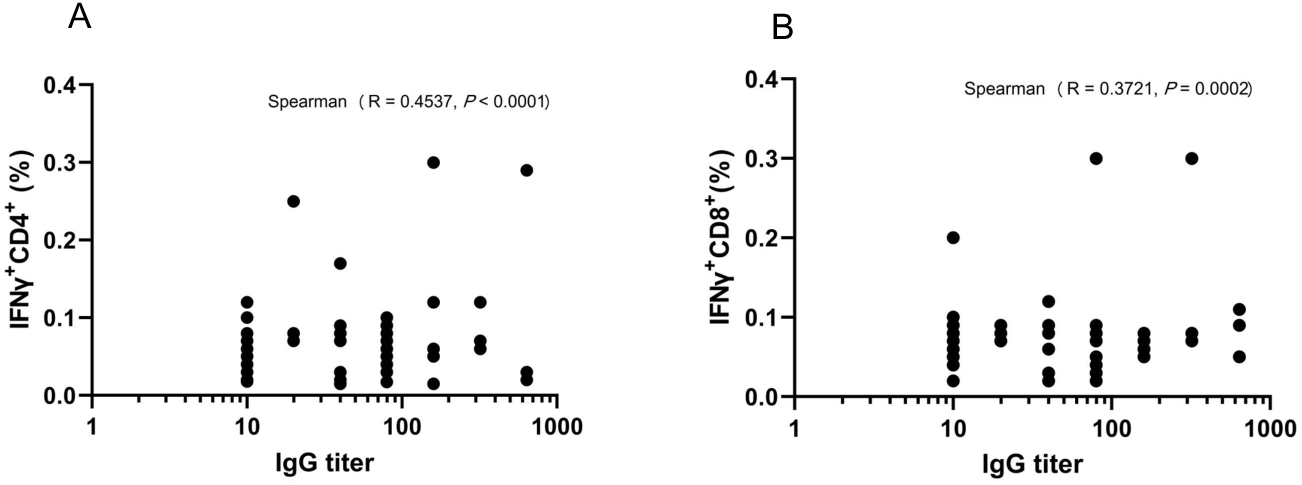
Severe acute respiratory syndrome coronavirus 2 (SARS-CoV-2)-specific T-cell immunity (interferon [IFN]y+CD8+ and IFNγ+CD4+) showing no correlation with SARS-CoV-2-specific humoral immunity.

**Figure 7.**
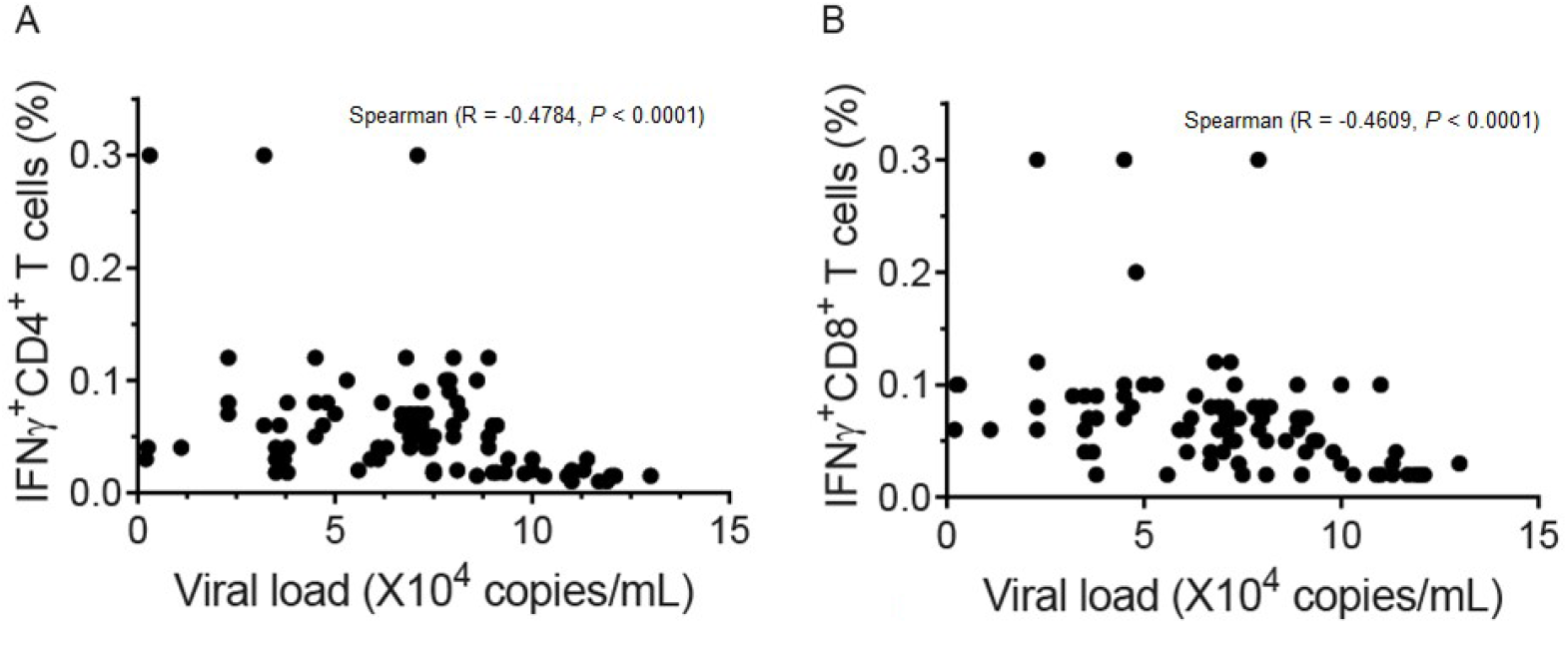
Inverse correlation of severe acute respiratory syndrome coronavirus 2 (SARS-CoV-2)-speciflc T-cell immunity (interferon [IFN]y+CD8+ and IFNγ+CD4+) with SARS-CoV-2 viral load.

SARS-CoV-2-specific humoral immunity inversely correlated significantly with SARS-CoV-2 viral load, which suggests that SARS-CoV-2-specific humoral immunity played a role in viral clearance (Figure 8). However, the R value of SARS-CoV-2-specific humoral immunity was smaller than that of SARS-CoV-2-specific T-cell immunity (Figures 7 and 8), which suggests that SARS-CoV-2-specific T-cell immunity might play a major role in viral clearance in fatal cases.

**Figure 8.**
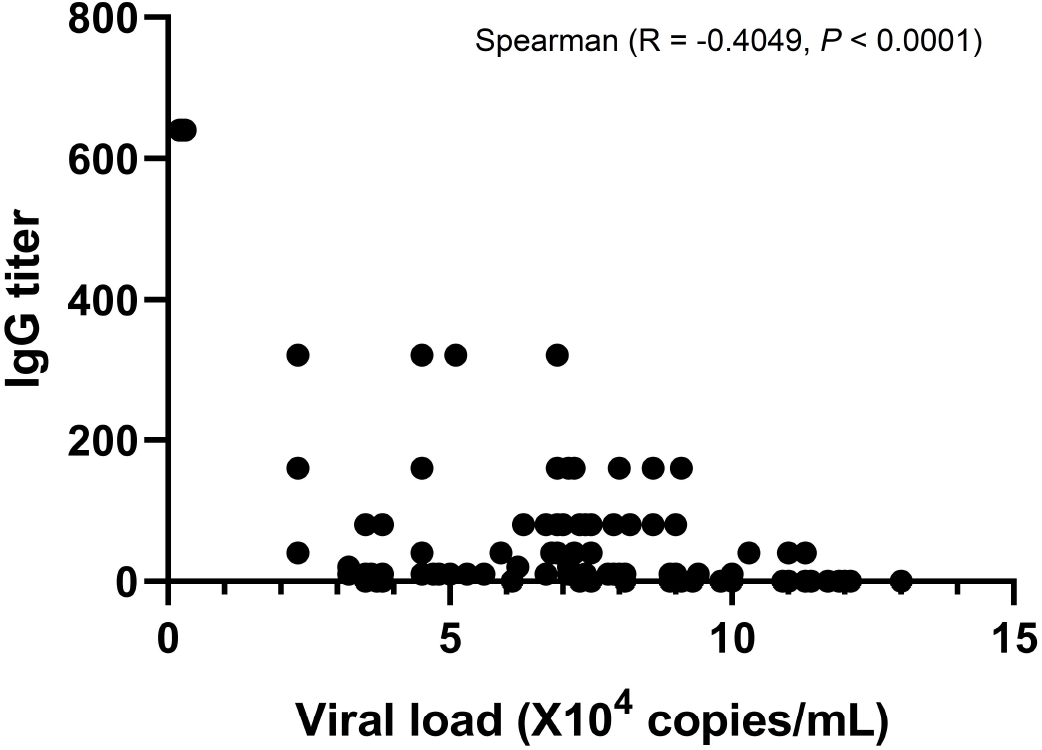
Inverse correlation of severe acute respiratory syndrome coronavirus 2 (SARS-CoV-2)-specific humoral immunity (IgG) with SARS-CoV-2 viral load.

However, the proportion of patients with SARS-CoV-2-specific T cells was significantly lower than that of COVID-19 survivors (Figure 9A, 9B), which suggests a loss in the expansion of the SARS-CoV-2-specific cellular immunity in fatal cases. In addition, the IgG titer was significantly lower than that in COVID-19 survivors (Figure 9C), which suggests a loss in the expansion of SARS-CoV-2-specific humoral immunity in the fatal cases. Here, we define "loss in the expansion" as a decrease in SARS-CoV-2-specific immunity in fatal cases compared to survivors of critical COVID-19 cases.

**Figure 9.**
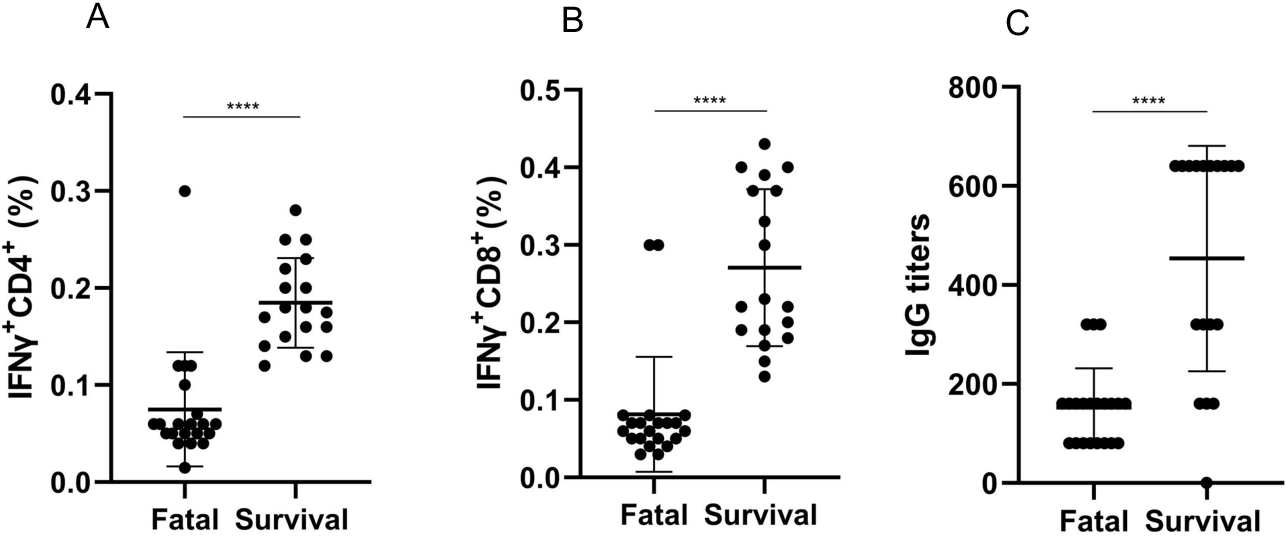
Comparison of severe acute respiratory syndrome coronavirus 2 (SARS-CoV-2)-specific immunity at week 3 between the 21 fatal cases and 18 survival cases of critical COVID-19. The graph showed the percentage of SARS-CoV-2 specific IFNγ+CD4+T cells (A), SARS-CoV-2 specific IFNγ+CD8+ T cells (B) and IgG titers (C). ****, *P* < 0.0001.

Furthermore, SARS-CoV-2-specific immunity was unable to clear the SARS-CoV-2 replications in the fatal cases. All the patients had a positive viral load before death, which suggests that prolonged SARS-CoV-2 exposure can predict fatal outcomes (Figure 4D).

## DISCUSSION

The extent of lymphopenia in patients admitted to the intensive care unit correlates with COVID-19 severity and mortality^23-26^. Therefore, studies of the immune system and function in healthy controls are important to understand whether the immune system and function is related to age. Our data showed that the immune system and function have gradually remodeled and declined with age in 25,239 healthy controls.

Channappanavar et al.^27^ reported that SARS-CoV-specific memory CD8 T cells persisted for up to 6 years after SARS-CoV infection. When challenge with a lethal dose of SARS-CoV, virus-specific memory CD8 T cells efficiently produced IFNγ, tumor necrosis factor α (TNF-α), etc. and reduced lung viral load. Next, we tried to understand whether SARS-CoV-2-specific immunity is associated with age in those fatal patients with COVID-19. Our data suggested that SARS-CoV-2-specific immunity has declined with age in COVID-19 patients.

Levels of lymphocytes and lymphocyte subsets are of great importance to keep the immune system working. Usually viral infection, immunodeficiency diseases, and other infectious diseases lead to abnormal changes in the levels of lymphocyte subsets^16^. Although SARS-CoV-2 has been identified as the causative agent of COVID-19, the mechanism by which SARS-CoV-2 impacts the human immune system is still unclear^16^.

Grifoni et al.^22^ reported SARS-CoV-2-specific CD4 and CD8 T cells responses in 20 recovery cases of COVID-19. SARS-CoV-2-specific CD4 T cell and antibody responses were observed in all COVID-19 patients and SARS-CoV-2-specific CD8 T cell responses were seen in most COVID-19 patients. Ni et al.^18^ also observed SARS-CoV-2-specific humoral and cellular immunity in 14 convalescent patients. We explored whether SARS-CoV-2-specific immunity is associated with survival time in those fatal patients with COVID-19. Our data showed that SARS-CoV-2 specific immunity was associated with survival time in COVID-19 patients. Our results were consistent with previous reports^18,22^.

From the above results, it can be known that the key factor was loss in the expansion of SARS-CoV-2-specific humoral and cellular immunity and the virus cannot be eliminated from the body in fatal cases. We tried to understand whether loss in the expansion of SARS-CoV-2-specific immunity in fatal cases could be expanded *in vitro*. Since the expansion of SARS-CoV-2-specific immunity started from the first 2-3 weeks, we chose PMBCs in the third week for the expansion experiment *in vitro*. Our data showed that these PBMCs could be expanded *in vitro*. After the expansion, SARS-CoV-2-specific IFNγ+CD4+ T cells and IFNγ+CD8+ T cells increased significantly. It suggested that the SARS-CoV-2-specific immunity of these fatal patients lost their expansion, but the expansion of SARS-CoV-2-specific cellular immunity could be recovered *in vitro* experiment.

Our data indicated that a concurrent decline in SARS-CoV-2 specific cellular and humoral immunity and prolonged SARS-CoV-2 exposure predicted fatal outcomes. First, the immune system and function have gradually remodeled and declined with age in healthy controls. Therefore, the elderly are susceptible to the SARS-CoV-2. Second, our data showed that SARS-CoV-2-specific immunity has declined with age in COVID-19 patients. Therefore, the elderly easily turned from mild to severe SARS-CoV-2 infections. Those results have explained that the elderly is a risk factor for poor outcomes. Third, our data further showed that SARS-CoV-2-specific immunity is associated with survival time in COVID-19 patients. The correlation between SARS-CoV-2-specific IFNγ+CD4+ T cells and IFNγ+CD8+ T cells and survival time was relatively strong, and significantly correlated with survival time, suggesting that patient survival time depended on the expansion ability of SARS-CoV-2-specific T cell immunity. The aggregated data for 4 weeks showed that SARS-CoV-2-specific humoral immunity was related to survival time. SARS-CoV-2 viral load was inversely proportional to survival time significantly. It suggested that the higher the viral load of SARS-CoV-2, the shorter the survival time of patients. Therefore, controlling the replication of the virus might prolong the survival time of patients. Finally, we observed that a concurrent decline in SARS-CoV-2-specific cellular and humoral immunity and prolonged SARS-CoV-2 exposure predicted fatal outcomes. There was significant correlation between SARS-CoV-2-specific T cell immunity and SARS-CoV-2-specific humoral immunity. The frequency of SARS-CoV-2-specific T cells and titer of IgG were much lower than that of survivors of critical COVID-19 cases and there were higher viral load in the course of the disease, indicating that the persistent SARS-CoV-2 infection and loss in the expansion of SARS-CoV-2-specific immunity are important factors.

## Data Availability

We have all data in the experiment.

## Acknowledgments

The work was supported in part by the National Natural Science Foundation of China (Grant No. 81830052 and 81530053).

## Author contributions

^#^Drs. Q Zeng, G Huang and YZ Li contributed equally to this article and share first authorship.

^*^Correspondence to Dr. Y Xu.

Q Zeng, G Huang and YZ Li contributed to study design, data interpretation, and manuscript drafting. GX, ZTC, ZTC contributed to data analysis, figure preparation, the literature search, and manuscript drafting. SYD and GX contributed to performed experiments, data collection, data analysis, and figure preparation. YX contributed to study design and reviewed the final draft. All authors read and approved the manuscript.

## Competing interests statement

We declare no competing interests.

## Supplemental Materials

**Table S1.** Demographic characteristics of 35 fatal patients with COVID-19 on admission.

| **Characteristics** | **Patients** |
| --- | --- |
| **Age, Median (IRQ), years** | 64.0 (45.0-73.0) |
| **Gender** |  |
| Male | 24 (68.6%) |
| Female | 11 (31.4%) |
| **Comorbidities** |  |
| Hypertension | 11 (31.4%) |
| Cardiovascular diseases | 7 (20.0%) |
| Diabetes | 6 (17.1%) |
| **Main symptoms** |  |
| Fever | 30 (85.7%) |
| Cough | 18 (51.4%) |
| Fatigue | 6 (17.1%) |

**Table S2.** Spearman’s correlation test and mean of lymphocyte subsets in 25,239 healthy controls.

| **Age** | **CD3<sup>+</sup>** | **CD4<sup>+</sup>** | **CD8<sup>+</sup>** | **CD4<sup>+</sup>/CD8<sup>+</sup>** | **CD19<sup>+</sup>** | **CD16<sup>+</sup>CD56<sup>+</sup>** |
| --- | --- | --- | --- | --- | --- | --- |
| year | cells/mm<sup>3</sup> | cells/mm<sup>3</sup> | cells/mm<sup>3</sup> | ratio | cells/mm<sup>3</sup> | cells/mm<sup>3</sup> |
| **16-19** | 1835 | 857 | 621 | 1.24 | 304 | 257 |
| **20-29** | 1648 | 876 | 594 | 1.49 | 279 | 180 |
| **30-39** | 1502 | 852 | 526 | 1.64 | 247 | 188 |
| **40-49** | 1417 | 829 | 465 | 1.81 | 238 | 189 |
| **50-59** | 1379 | 822 | 428 | 1.92 | 237 | 200 |
| **60-69** | 1309 | 763 | 421 | 1.84 | 230 | 244 |
| **70-79** | 1220 | 718 | 391 | 1.82 | 210 | 265 |
| **80-91** | 938 | 530 | 348 | 2.09 | 166 | 304 |
| **R** | -1 | -0.9762 | -1 | 0.9048 | -1 | 0.6429 |
| **P** | <0.0001 | 0.0004 | <0.0001 | 0.0046 | <0.0001 | 0.0962 |

**Figure S1.**
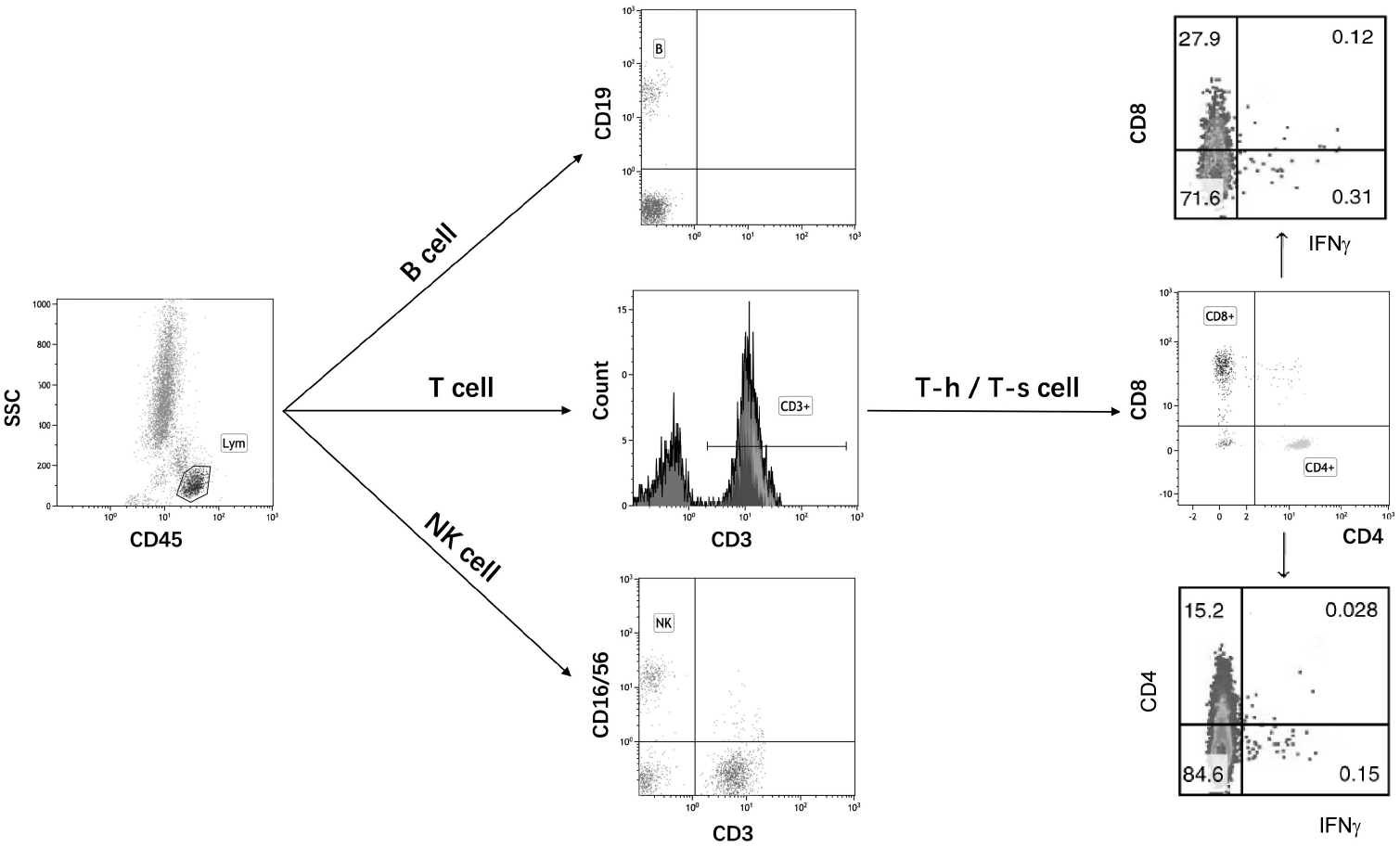
A representative flow cytometry gating strategy.

## EXPERIMENTAL MODEL AND SUBJECT DETAILS

### Study Design and Participants

This retrospective study was conducted at Chinese PLA General Hospital, Peking Union Medical College Hospital, the First Affiliated Hospital, and affiliated hospitals of Shanghai University of Medicine and Health Sciences. Chinese PLA General Hospital and Peking Union Medical College Hospital are the largest clinical research hospitals in China. Demographic characteristics of 35 fatal patients with COVID-19 on admission were listed in Table S1. The 18 critical cases (matched by age and sex) who survived were enrolled for comparison. This case series was approved by the institutional ethics board of Chinese PLA General Hospital (#2020-111), Peking Union Medical College Hospital (#ZS-1830), The First Affiliated Hospital (#KY2020046), and Shanghai University of Medicine and Health Sciences (#2019-LCHZ-18-20190507). Written informed consent was obtained from healthy controls and waived to COVID-19 patients due to the rapid emergence of this infectious disease. Identification of hypertensive patients was achieved by reviewing and analyzing available electronic medical records and patient care resources. Clinical outcomes (discharge, mortality, and recovery, etc.) were monitored and all clinical recovery hypertensive patients meet the following criteria: body temperature returned to normal for more than 3 days, respiratory symptoms improved significantly, and lung imaging showed significant improvement.

## METHOD DETAILS

### Cell preparation

Whole blood was centrifuged for 15 min at 1800 rpm to separate the cellular fraction and plasma. The plasma was then carefully removed from the cell pellet and stored at −20°C. Peripheral blood mononuclear cells (PBMCs) were isolated using Ficoll-Paque PLUS density gradients (GE Healthcare Life Sciences, USA) according to the manufacturer’s instructions. Isolated PBMCs were either studied directly or cryopreserved in cell recovery media containing 10% DMSO (Gibco, USA), supplemented with 10% heat inactivated fetal bovine serum and stored in liquid nitrogen until used in the assays. Cryopreserved PBMCs were thawed by diluting them in 10 mL complete RPMI 1640 with 5% human AB serum (Gemini Bioproducts, USA) in the presence of benzonase (final concentration at 50 U/mL) before an experiment.

### SARS-CoV-2 Spike glycoprotein peptide pools

SARS-CoV-2 Spike glycoprotein peptide pools (SPs) were from Genscript Biotech, USA. SPs include 316 peptides (delivered in two subpools of 158 peptides) derived from a peptide scan (15 mers with 11 amino acid overlap) through the entire Spike glycoprotein of SARS-CoV-2.

### Expansion of PBMCs *in vitro*

20% of PBMCs were pulsed with 10 μg/mL of the SPs for 1 hour at 37 °C, subsequently washed, and cocultured with the remaining cells in AIM-V medium (Gibco; Thermo Fisher Scientific, USA) supplemented with 2% AB human serum (Gibco; Thermo Fisher Scientific, USA). PBMCs were cultured for 7 days in the presence of 20 U/mL of recombinant IL-2 (R&D Systems, USA).

### Intracellular cytokine staining (ICS) assay

PBMCs were first stimulated with or without SPs (2 μg /mL) for 3 h at 37°C, then brefeldin A (10 μg/mL, Sigma-Aldrich, USA) was added to cultures to enable intracellular proteins to accumulate in all stimulations. PBMCs were stimulated RPMI medium containing 10% FCS as a negative control. After incubation for a total of 6 h, the cells were washed, fixed, permeabilized using fixation/permeablization solution kit (BD Biosciences, USA) and blocked with FcR blocking reagent (Meltenyi Biotec, USA) for 30 min at 4 °C to reduce nonspecific binding of antibodies to human Fc receptor. The cells were then stained with anti-IFNγ antibodies (BD Bioscience, USA) for 30 min at 4 °C. After staining, all samples were washed twice with phosphate buffered saline (PBS) containing 0.1% saponin, 0.1% BSA and 0.05% NaN_3_ (Sigma-Aldrich, USA), and resuspended in 300 μL PBS for measurement in a flow cytometer (FACSVerse™ flow cytometry, BD Bioscience, USA).

### Real-time reverse transcription polymerase chain reaction (RT-PCR) assay for SARS-CoV-2

Throat, sputum and blood samples were collected for extracting SARS-CoV-2 RNA from hypertensive patients suspected of having SARS-CoV-2 infection. In brief, RNA was extracted within 2 h using the total RNA isolation kit. Cell lysates (250 μL) were transferred into a collection tube followed by vortex for 10 s. After standing at room temperature for 10 min, the collection tube was centrifugated at 1000 rpm for 5 min. The suspension was used for RT-PCR assay of SARS-CoV-2 RNA. Two target genes, including open reading frame 1ab (ORF1ab) and nucleocapsid protein (N), were simultaneously amplified and tested during the RT-PCR assay. The sequences of primers and probes were: Target 1 (ORF1ab): forward primer CCCTGTGGGTTTTACACTTAA; reverse primer ACGATTGTGCATCAGCTGA; and the probe 5ʹ-VIC-CCGTCTGCGGTATGTGGAAAGGTTATGG-BHQ1-3ʹ. Target 2 (N): forward primer GGGGAACTTCTCCTGCTAGAAT; reverse primer CAGACATTTTGCTCTCAAGCTG; and the probe 5ʹ-FAM-TTGCTGCTGCTTGACAGATT-TAMRA-3ʹ. The RT-PCR assay was performed using a SARS-CoV-2 nucleic acid detection kit. Reaction mixture contained 12 μL of reaction buffer, 4 μL of enzyme solution, 4 μL of probe primer solution, 3 μL of diethyl pyrocarbonate-treated water, and 2 μL of RNA template. The RT-PCR assay was performed under the following conditions: incubation at 50 °C for 15 min and 95 °C for 5 min, 40 cycles of denaturation at 94 °C for 15 s, and extending and collecting fluorescence signal at 55 °C for 45 s. A cycle threshold value (C_t_-value) less than 37 was defined as a positive test result, and a C_t_-value of 40 or more was defined as a negative test. These diagnostic criteria were based on the recommendation by the National Institute for Viral Disease Control and Prevention (China). Samples with a C_t_-value of 37 to less than 40 was confirmation by retesting. The copies of RNA per reaction were obtained from the standard curve of limiting-dilution series of standard copies of RNA versus PCR amplification cycle.

### Enzyme-linked immunosorbent assay (ELISA) of immunoglobulin (Ig)

Antibodies (IgM, IgA, and IgG) specific to SARS-CoV-2 were determined with two different ELISA: an in-house assay using SARS-CoV-2 Receptor Binding Domain (RBD) protein (Cat. #Z03479, Genscript Biotech, USA) as an antigen, or a commercial kit (SARS-CoV-2 Spike RBD ELISA Kit, Cat. #40591-V08H, Sino Biological, China). Microtiter plates were coated with 50 ng/well of target protein overnight at 4 °C. Plates were then blocked for 2 h at 37 °C using 200 μL of 5% non-fat milk in PBS. Serum samples were then diluted into 1:50 using PBS and 100 μL of each sample was applied to coated ELISA plate and incubated for 2 h at 37 °C. Plates were then washed with PBS and incubated with HRP-labeled anti-human IgM, IgA, and IgG (Sigma Aldrich, USA), which were diluted to 1:2000 in 5% non-fat milk in PBS. After incubation for another 1 h at room temperature, the plates were washed and developed with TMB/E substrate (Millipore, USA). Finally, the reaction was stopped with 1 M H_2_SO_4_, and the optical density (OD) at 450 nm was measured. Negative serum control was run each time when the assay was performed. A sample is positive if its adjusted OD value (OD of test - OD of control) exceeds the mean plus 3 standard deviations of the normal controls.

### Flow cytometry analysis

All samples were analyzed by FACSVerse™ flow cytometry (BD Bioscience, USA). Two blood samples in two tubes (100 μL each) were stained according to the manufacturer’s instructions. Then, red-cell lysis buffer (1 mL) was added to each tube, the samples were incubated for 10 min and washed with Sorvall cell washer (Thermo Fisher Scientific, USA). Cells were blocked with FcR blocking reagent (Meltenyi Biotec, USA) for 30 min at 4 °C to reduce nonspecific binding of antibodies to human Fc receptor and washed with PBS. The cells were then stained with antibodies as listed in Figure and the fixable dead cell stain kit (Invitrogen, USA) for 30 min at 4 °C and were then resuspended in 350 μL PBS and analyzed in flow cytometry (FACSVerse™ flow cytometry, BD Bioscience, USA). Calibration and quality control for the instrument were carried out daily with the use of eight-color setup beads (BD Bioscience). All specimens were analyzed in duplicates with coefficient of variation (CV) < 5% by two independent technicians under the inter-laboratory quality control. The experiments were repeated if the results showed CV > 5% according to the instructions of BD Bioscience. The data were analyzed by FlowJo software (version 10, Tree Star, USA).

### Statistical analysis

Categorical variables were described as frequency rates and percentages, and continuous variables were described using mean and median values. Means for continuous variables were compared using independent group *t* tests when the data were normally distributed; otherwise, the Mann-Whitney test was used. Data (non-normal distribution) from repeated measures were compared using the generalized linear mixed model. Proportions for categorical variables were compared using the χ^2^ test, although the Fisher’s exact test was used when the data were limited. All statistical analyses were performed using SPSS (Statistical Package for the Social Sciences Inc., version 13.0). For unadjusted comparisons, a two-sided *P* value less than 0.05 was considered statistically significant. The analyses have not been adjusted for multiple comparisons and, given the potential for type I error, the findings should be interpreted as exploratory and descriptive. Because the cohort of patients in our study was not derived from random selection, all statistics are deemed to be descriptive only.

## Notes

### Competing Interest Statement

The authors have declared no competing interest.

### Author Declarations

This case series was approved by the institutional ethics board

